# Opportunities for preconception care in Australia: A qualitative study of the perspectives of primary care professionals

**DOI:** 10.1101/2025.01.16.25320705

**Authors:** Cherie Caut, Danielle Schoenaker, Erica McIntyre, Amie Steel

## Abstract

**Background:** Preconception care is well situated within primary care settings where pregnancy intention and preconception risk screening can occur routinely, along with coordinating preconception health interventions. Implementing preconception care into primary care practices has been challenging, as health professionals report multiple barriers preventing effective preconception care. A clearer insight into the scope of preconception care within Australian primary care settings is needed. Therefore, this study aimed to explore the beliefs and attitudes towards preconception care and describe the preconception care practices of general practitioners, midwives, and naturopaths in Australia to identify opportunities for preconception care.

**Methods:** This study employed a qualitative methodology involving interviews and focus groups with primary care health professionals (n=18) in clinical practice (> 5 years) within Australia. General practitioners (GPs) (n=6), midwives (n=5), and naturopaths (n=7) were recruited through professional organisations between May and August 2021. The fieldwork explored participants’ beliefs and attitudes towards preconception care and their preconception care practice behaviours. A thematic framework approach applicable to health services research was employed for data analysis.

**Results:** The analysis identified three key themes about the participants’ perspectives on providing preconception care: *Preconception health requires a population-level focus across the life stages (subthemes: public health focus on preconception health and care is insufficient*, *more public information on preconception health and care is needed*, and *public education on preconception health and care requires a life course approach), Men are being overlooked and women carry the burden of responsibility in preconception care (subthemes: men lack awareness of their role in preconception care, men are not usually a part of preconception care consultations, men have unique needs and require different approaches to engage in preconception care*, and *women are carrying the burden of responsibility in preconception care),* and *Coordination of preconception care among primary care providers (subthemes: GPs as primary care providers of preconception care, health professionals other than GPs as providers of preconception care*, and *health professionals other than GPs want greater coordination of preconception care)*.

**Conclusions:** Health professionals have identified that making the public more aware of preconception health and aligning clinical care and public health initiatives are important. Health professionals should think more carefully about engaging with men in preconception care. Encouraging men to participate in preconception care requires different information and approaches. While GPs are positioned in the healthcare system as responsible for the coordination of care, there are potential contributions to preconception care that other health professionals can make due to their position in the healthcare system, their skills and the populations they service. A complementary and collaborative whole healthcare system approach to preconception care is needed and may provide more opportunities to deliver preconception care to improve reproductive outcomes.

## Introduction

Preconception care is well situated within primary care settings where pregnancy intention and preconception risk screening can occur opportunistically or routinely, along with coordinating preconception interventions [1, 2]. The World Health Organisation defines primary healthcare as “a whole-of-society approach to effectively organise and strengthen national systems to bring services for health and wellbeing closer to communities [3].” Preconception care comprises screening for preconception risks and providing preventive, promotive, curative health, and social intervention [4, 5] to reduce maternal and childhood mortality and morbidity [5]. Concerning both the maternal and paternal reproductive partner [6, 7], multiple modifiable and non-modifiable preconception risks and health behaviours exist (e.g., body composition, lifestyle, nutrition, environmental exposures, birth spacing, maternal age, genetic disorders, chronic and infectious diseases, medications, mental health conditions and intimate partner violence) [6, 8].

A range of primary care health professionals have opportunities to provide preconception health assessment, education, and counselling [9] in any clinical or hospital setting [10] before a first pregnancy and between pregnancies [4, 5]. For example, women consult with general practitioners (GPs) and midwives as providers of preconception care [1]. GPs have opportunities for preconception care as they regularly have contact with women of reproductive age and the general population[2]. Midwives have contact with women between their pregnancies, provide health promotion as a part of their role [11, 12] and have opportunities to provide preconception care between women’s pregnancies (i.e., to provide interconception care) [2, 13]. Naturopaths are more likely to be accessed by women who are attempting to conceive than women who are not [14]. Also, as providers of health promotion [15], naturopaths have opportunities to provide preconception health information before pregnancy [16].

Despite the opportunities for providing preconception care in primary care health settings, health professionals have found that implementing effective preconception care into their practices has been challenging as they are often met with barriers. Health professionals have reported that the barriers to preconception care include limited time, practitioner or patient knowledge gaps, and a lack of referral and coordination between health professionals [11, 17-20]. Furthermore, health professionals believe that preconception care can be enabled by patients who raise the topic with their clinician, either directly because they are aware of the benefits or indirectly through mentioning an intention to become pregnant [11, 18-20].

A clearer insight into the scope of preconception care within Australian primary care settings is needed. This insight will help inform Australian healthcare policymakers and healthcare service administrators of opportunities for the nationwide implementation of effective preconception care services. Therefore, this study aimed to explore the beliefs and attitudes towards preconception care and describe the preconception care practices of general practitioners, midwives, and naturopaths in Australia to identify opportunities for preconception care.

## Methods

### Research design

The study design used a phenomenological theoretical framework [21]. A qualitative methodology explored the complex phenomena health professionals encounter in preconception care [21]. Data collection methods comprised semi-structured interviews and focus groups [21]. The ‘Consolidated criteria for reporting qualitative research (COREQ): a 32-item checklist for interviews and focus group’ [21] was used to report the data results descriptively.

### Participants

GPs, midwives, and naturopaths in Australia who have worked in clinical practice for at least five years were invited to attend an online focus group or interviews.

### Setting

The Zoom Video Communications platform was used to facilitate and audio record interviews and focus groups for participants attending from a range of geographical locations within Australia, including remote regions, of which one of each interview and focus groups were held for GPs and midwives and two focus groups held for naturopaths. Two research team members (AS and CC) facilitated each focus group, and one member (CC) facilitated the interviews. The duration of each interview and focus group were 30 to 40 minutes and 80 to 90 minutes, respectively.

### Recruitment

Professional organisations, including RACGP (Royal Australian College of General Practitioners), RACGP Specific Interest group Antenatal and Postnatal Care, Australian College of Midwives (ACM), Naturopaths and Herbalists Association of Australia (NHAA) and Complementary Medicines Association (CMA) provided the channels for participant recruitment via direct email, newsletter, or social media. A gift voucher incentive of AUD 50.00 for midwives and naturopaths and $100.00 for GPs was offered to encourage participation in our study. This figure was selected based on input from stakeholder groups (RACGP), published data about practitioner hourly fees in clinical practice [22] and ensuring equality between groups [23]. Interested health professionals could view the Participant Information Sheet and eligibility criteria and provide their consent to participate in the study via a short screening survey developed in Qualtrics XM™ online survey software [24]. The screening survey collected respondents’ preferred availability and contact information to facilitate the organisation of the focus groups and interviews by members (CC, AS) of the research team. The selection of participants occurred through purposive sampling methods [21]. An email invitation to participate was sent out to respondents, stating the day and time preferred by most respondents. The phases of recruitment and retention of participants in the study are detailed in Figure 1.

**Figure. 1.**
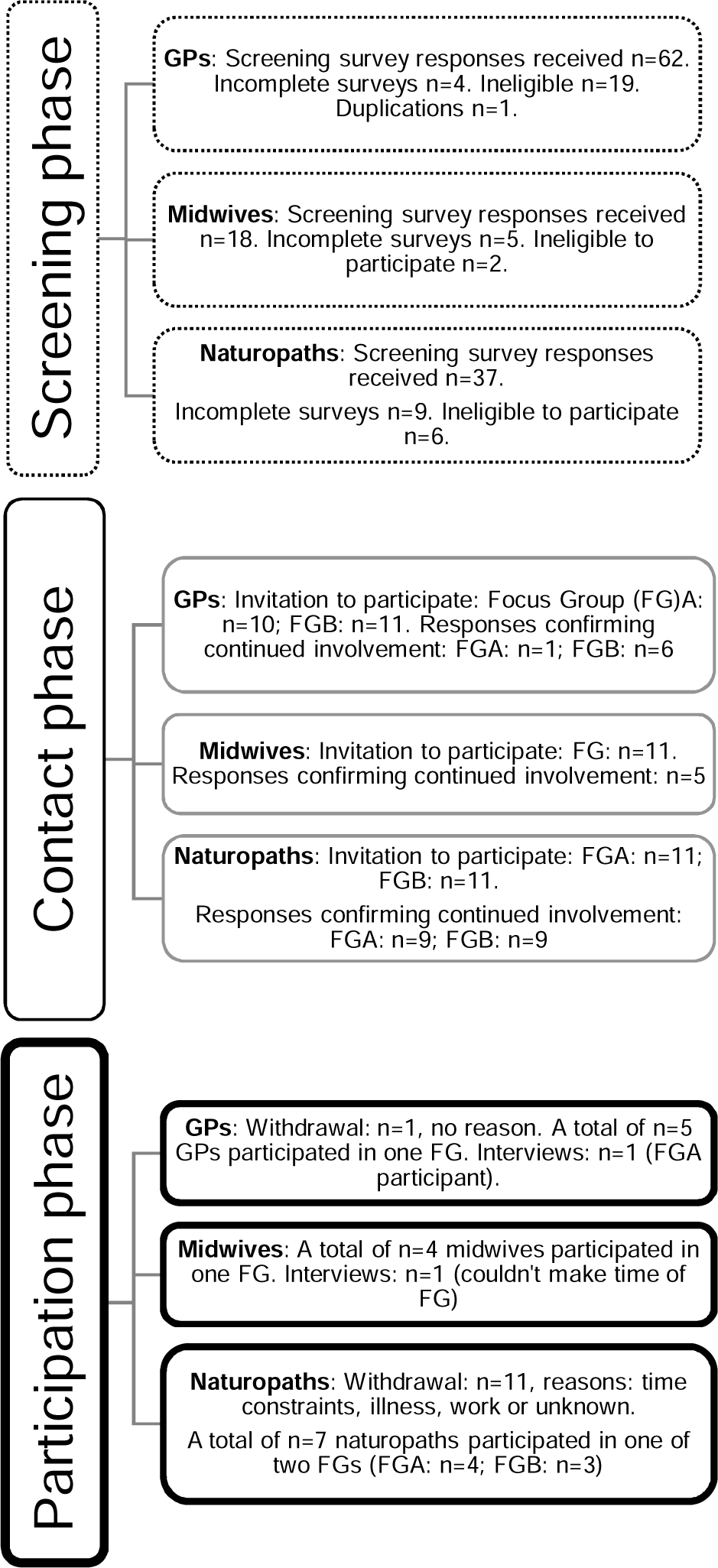
Phases of participant recruitment and retention.

### Sample

Between May and August 2021, eighteen health professionals in current clinical practice (> 5 years) within Australia participated in online focus groups and interviews comprising GPs (n=6), midwives (n=5), and naturopaths (n=7). One interview and focus were held for GPs and midwives respectively, and two focus groups for naturopaths. The number of participants in each focus group ranged between three to five (GPs: [n=1 focus group; n=5 participants], midwives: [n=1 focus group; n=4 participants], naturopaths [n=2; focus group A, n=4 participants; focus group B, n=3 participants]). The duration of each interview and focus group were 30 to 40 minutes and 80 to 90 minutes, respectively.

### Data collection

Participants were invited to share their perspectives and experiences as health professionals providing preconception care during the interviews and focus groups. A focus group and interview guide were developed to facilitate discussion and guide the interview (see Additional file 1) and explore domains: practice services provided, beliefs and attitudes towards preconception care, and preconception care practice behaviours. At n=16 focus group participants, thematic saturation was achieved, and the interviews of one each for GPs and midwives facilitated surety of this.

### Data analysis

The audio recordings from the interviews and focus groups were transcribed by Rev audio transcription service [25] and imported into NVivo 12 Pro qualitative data analysis program software [26] to facilitate part of the analysis process. Using a framework thematic analysis approach [27], data were analysed from transcripts by two research team members (CC, AS). The framework approach is an established method of analysis within health service research employing five stages: familiarisation, identifying a thematic framework, indexing, charting, and mapping and interpretation [27]. Quotes were selected based on the representativeness of the theme and the quality of the quote.

## Results

Descriptive thematic analysis of participants’ accounts of providing preconception care in Australian primary health care settings identified shared experiences and perspectives across all health professions and other experiences described by participants from one or two health professions. Three key themes were identified across the explored domains: beliefs and attitudes towards preconception care and practice behaviours, categorised as *Preconception health requires a population-level focus across the life stages, Men are being overlooked and women carry the burden of responsibility in preconception care,* and *Coordination of preconception care among primary care providers*.

### Preconception health requires a population-level focus across the life stages

The first theme covered participants’ views concerning the current level of public health, focusing on preconception health. One subtheme addressed that the ‘public health focus on preconception health and care is insufficient,’ and the second addressed that ‘more public information on preconception health and care is needed.’ The third subtheme addressed that the ‘public education on preconception health and care requires a life course approach.’

### Public health focus on preconception health and care is insufficient

Health professionals in our study expressed a shared view that the current level of public information available on preconception health or care was insufficient, as evident in these quotes from a naturopath, GP and midwife participant:

> *“The public level of information or the public level of understanding about how much our lifestyles and exposures affect our fertility is fairly low.” (Naturopath 7)*

> *“I don’t think there’s very much. I don’t remember ever as a lay person seeing any information anywhere on preconception care, unless you go to the pharmacy and then you might notice pregnancy vitamins, but otherwise, I haven’t seen anything on social media or on TV, you just don’t see anything.” (GP 6)*

> *“Well, I wonder who is offering preconception information or education here…So there’s not a lot available it seems.” (Midwife 5)*

Although participants across the health professional groups agreed that there was some evidence of increasing attention towards preconception health in public health as expressed by this naturopath describing some examples of the available preconception health information initiatives including health promotion websites (e.g. YourFertility) and clinical services (e.g., IVF group):

> *“There’s various people trying; Your Fertility, the IVF group, put out the basics on weight, age, lifestyle issues, and they’re trying to reach the GPs with that information as far as I understand. So, there are groups that are trying to get this across, but…I just think, at the moment there really is very little. A lot of patients are really not aware of a lot of this information when they walk through the door.” (Naturopath 4)*

### More public information on preconception health and care is needed

Health professionals in our study shared their views on the information available on preconception health. One GP participant described how it is most likely that patients would search the internet for preconception health information yet raised their concern for the potential lack of that information:

> *“I guess patients would probably just Google if they are looking for that information. And then who knows what they’ll find. See, I don’t think there’s a lot of public health messaging around it.” (GP 6)*

This naturopath participant described their belief that a greater focus on public health information would be helpful and how an increase in public preconception health education would support those health providers providing preconception care by bridging the knowledge gap on the associations between preconception health and maternal and child life course health outcomes:

> *“It’d be lovely if it was more of a public health thing because I guess the issue I come up against a lot is that it feels like there’s a bit of an uphill battle often to get women on the same page, to get people to the point where they understand the importance of what we’re trying to do from a naturopathic perspective, to get their health to a point where they can have a healthy baby, because I think naturopaths tend to be looking at things from the perspective of the longer term outcome rather than that positive pregnancy test… Because that information is not very well understood in the general community, I think that makes the preconception health care more difficult.” (Naturopath 7)*

This GP expressed concern that current public health messages towards women concerning preconception health can tend to be negative:

> *“I almost feel sometimes we scare our female population about thinking about this [preconception health] or talking about this [preconception health], because… sometimes I think it’s [preconception health] in the negative sort of connotation.” (GP 1)*

### Public education on preconception health and care requires a life course approach

The health professionals in this study shared strong views concerning the ideal place and timing for educating the public on preconception health. Health professionals agreed that preconception health education should be within schools and commence during adolescence, as captured in these quotes:

> *“Perhaps it starts in the schools. Again, like in the high school setting. So whether it is … when upskilling even like school chaplains and things like that.” (Midwife 3)*

> *We do very well at contraception, and I think we do that very well from when we’re in school…so it would be nice to have a little bit more of a balanced view, maybe, from that perspective, especially maybe coming into young adulthood where this [preconception health] is something. (GP 1)*

> *“I think the behaviours of people, as I said, right from puberty through and even before that really, are going to have an impact on their fertile health, but it’s not a conversation that’s had anywhere that I can say that I’m aware of.” (Naturopath 7)*

Health professionals shared their beliefs about the ideal time frame for commencing preconception care, all agreeing that preconception care should be considered before pregnancy. Naturopath participants believe that preconception care is best commenced three to four months before trying to conceive, as described by this naturopath:

> *“We do our best to persuade them to stop trying while we complete three to four months of preconception healthcare.” (Naturopath 1)*

GP participants also expressed the view that preconception care is best delivered as early as possible, with one GP indicating it can benefit women even when they are single:

> *“I think the earlier the better. I think if a woman, if she’s not even in a relationship…that the earlier the better, and I think that the more education that women have with regards to this, then they can really start making some decisions on their own.” (GP 5)*

Midwives’ participants shared their view that preconception care could be commenced anytime throughout the reproductive age when there is a possibility for pregnancy, as described by this midwife:

> *“Just opening up that conversation [pregnancy intention screening] in your primary healthcare setting all the time.” (Midwife 4)*

### Men are being overlooked and women carry the burden of responsibility in preconception care

The second theme identified covered participants’ views concerning men’s and women’s roles in preconception care. One subtheme addressed that ‘men lack awareness of their role in preconception care,’ and the second addressed that ‘men are not usually a part of preconception care consultations.’ A third subtheme addressed that ‘men have unique needs and require different approaches to engage in preconception care,’ the fourth subtheme addressed that ‘women are carrying the burden of responsibility in preconception care.’

### Men lack awareness of their role in preconception care

Shared among all health professional groups in our study were strong beliefs about the role of men in preconception care. One reason proposed by the participants was that men themselves are not aware of the effect their health has on reproductive outcomes as captured by these health professionals:

> *“I feel like they’ve [men] never heard of it before. So, I do think a lot of it is new information to them. I think I’ve noticed with men they’re often surprised that alcohol has anything to do with sperm quality.” (GP 6)*

> *“And it’s about the men don’t necessarily realise that they have a role to play here, and that there’s things they can do.” (Midwife 1)*

> *“They [men] don’t understand, they didn’t know the full role of that preconception period and the importance of it, and the importance of their health in that.” (Naturopath 4)*

GPs and midwives raised their concern that current public health messages on preconception health and care burden women with the responsibility and should include men more, as described in these quotes from a GP and a midwife participant:

> *“We get sort of taught that everything is about female’s responsibility when it comes to conception, contraception, anything to do with having children. We don’t think enough about dads, I think, either in terms of support or education or whatever.” (GP 4)*

> *“That [the role of men in preconception health] needs to be mainstreamed more, because we put a lot on women because they do come to health services.” (Midwife 1)*

### Men are not usually a part of preconception care consultations

One dominant view expressed by participants was that men are not usually a part of preconception care. Health professionals in our study described that women are usually the first members of a couple to make an appointment for preconception care and may often attend without their male reproductive partner, as described in these quotes from a GP and naturopath participant:

> *“I try my best to, at least the second consult, I try to encourage - and it’s usually the female partner that’s come in and started it - just to say, “Bring your partner along.” (GP 5)*

> *“We do get a lot of just women booking in.” (Naturopath 4)*

This midwife participant described the difficulty in getting men to engage in preconception care consultations:

> *“It’s hard to get those men in.” (Midwife 1)*

In response to being asked about the preconception care they provide to men, this GP participant described how they rarely consider men when it comes to preconception care and that becoming aware of the importance of their role during focus group discussions has caused them to want to improve the preconception care they provide to include men:

> *“I have to say I’m really bad with preconception care for men, I rarely think of it. I get less opportunities to do it opportunistically and then even when I do get the opportunities, I just don’t think I think of it. I think I should be talking about it; I think I should be talking about conception with men as well, obviously it’s equally their responsibility. I think I should do more, but I just don’t. Which is probably something worth reflecting on and trying to improve.” (GP 4)*

The GP and naturopaths in this study agreed that men are more motivated to participate in preconception care when a male infertility factor has been identified as suggested by this GP and naturopath:

> *“Men don’t come in for preconception counselling, until there’s a problem. I just don’t get a lot of men coming in to ask for preconception counselling until there’s a problem with the conception, then they come in to ask questions.” (GP 2)*

> *“I’ve also had men come to see me. Men that have had low sperm counts and they come with their wives and it’s become like the family wants to do this…sometimes, it’s the man that says I’ve got a low sperm count, what can I do to improve that, to improve my health so that we can have a baby naturally without having to do the IVF thing?” (Naturopath 6)*

### Men have unique needs and require different approaches to engage in preconception care

Another reason described by the participants for men not being engaged in preconception care is a perception that they require a different approach and type of information than women. These GPs described their preference to encourage men to general health care as opposed to preconception care and found this helpful in overcoming some of the hesitancy surrounding reproductive health checks:

> *“I try to sell it more as a general health check. I was like, “well, you’re looking to have a baby, you both want to be healthy” and that sort of thing, because a lot of them there is a bit of resistance in that, “there’s nothing wrong with me” it’s not suggesting, it’s just a general, “let’s see what your cholesterol is doing” those sort of factors. I just try to put it as a bit more of a broad health check” (GP 5)*

> *“Because you have that first visit with the woman on her own usually, you usually develop that rapport. And then the woman goes home and says, “Well, I really want you to come to the doctor. I’m so excited.” And then the man comes kind of a bit begrudgingly, but then when they come and realize it’s not that scary and we’re just having a chat and it’s nothing intrusive.” (GP 6)*

This naturopath participant described how the type of information provided to men regarding preconception care was better received when statistics, graphics and evidence-based information sources were used:

> *“It’s a little bit stereotypical and it’s certainly not always the case, but addressing the man with a lot more facts and figures certainly can get them onboard, and saying there is a section at each end of our chapter for him and that it’s all evidence-based, and the research is there if they want to look at it. That is a helpful approach.” (Naturopath 1)*

This GP participant shared their perception that men are generally less likely to seek health care, and this could be another barrier for men attending for preconception care:

> *“Also, when I’m talking to [a] woman who, for preconception stuff and I say I would like to see [the] man, it’s often quite difficult to get them into a clinic because they don’t have a GP, they’re not registered at our practice, they don’t see anyone. So then there’s a few barriers there, they’re just not in the habit of seeking health care.” (GP 4)*

### Women are carrying the burden of responsibility in preconception care

The health professionals in this study agreed that wherever possible, men are invited or encouraged to preconception care. Usually, however, the female partner is burdened with the responsibility to extend the invitation to attend a preconception care consultation or pass on the preconception health information to their male reproductive partner as described by these participants:

> *“I guess, for me, it’s mainly women that I see, but when they’re coming for preconception counselling, I often do encourage them to bring their male partner as well for him to also have that discussion or for him to see his regular doctor for that discussion.” (GP 6)*

> *“If the man is not amenable to coming in, we do try and address the man through her, but that is more tricky, but with diet and lifestyle, that’s possible, and certain basic nutrients, if they’re on a multi … But it is more tricky to get the men fully engaged, for sure.” (Naturopath 4)*

> *“And I’ll talk about those basic lifestyle things, and I usually will send her, at the end of the appointment,…my sperm health handout that talks about those lifestyle factors…so that at least I’ve got that information on paper for him to read and hopefully, he doesn’t have to front up and talk to me but he can read it and make some changes.” (Naturopath 7)*

> *“I do worry that either it [preconception health information] gets lost in translation, or that the women will only tell the partners a filtered version.” (Midwife 4)*

This midwife described that one of the reasons women are coming to preconception care and not men is due to the public perception that preconception care is a women’s health issue. Furthermore, this midwife described how their profession is predominantly women, which means there is a lack of men to discuss preconception health, especially adolescent boys who may feel embarrassed when the health professional discussing preconception health is a woman:

> *“I do think we land a lot of stuff on the women. And certainly, the context that I work in, it’s very much seen as women’s things. So, there’s not a lot of responsibility… very much all women stuff. The boys get very embarrassed…We do need male workers who can talk to the boys and can do the school stuff. And we know nursing…is a predominantly female led profession and so are midwives. But we do need the men to talk to the boys from 10 and 11.” (Midwife 1)*

### Coordination of preconception care among primary care providers

The third theme identified covered participants’ views concerning the coordination of care among primary care providers of preconception care. One subtheme addressed ‘GPs as primary care providers of preconception care’, and the second addressed ‘health professionals other than GPs as providers of preconception care.’ The third subtheme addressed that ‘health professionals other than GPs want greater coordination of preconception care.’

### GPs as primary care providers of preconception care

A dominant view expressed by naturopaths in our study was how they were aware of their own skill set, while emphasising the importance of the GPs’ role in providing effective preconception care. Naturopaths expressed their desire to share care with GPs when providing preconception care. Naturopaths described establishing referrals for their patients for preconception care to GPs, whether that be the patient’s existing GP or another:

> *“One of the things I always ask them [my patients] is, who is your GP? Because I always explain that I can do a certain amount, but I can’t do everything, and it’s important for them [my patients] to have a good GP on board.” (Naturopath 5)*

> *“I do have a doctor down the road that I refer to if someone doesn’t have a GP as well.” (Naturopath 6)*

This GP described how they did not think there was another health professional that could take on primary responsibility for providing preconception care as a GP can:

> *“If I was to think about who else would be able to take on that role [provide preconception care], I just don’t think there’s any other group of people.” (GP 1)*

### Health professionals other than GPs as providers of preconception care

The naturopaths in our study shared their belief that they, too, should be referred to for preconception care as they are skilled in providing preconception care and able to refer to other healthcare providers when required, as this naturopath described:

> *“I think in an ideal health environment, women should be referred to naturopaths for preconception care. I think we’re best placed to provide that care. And I off the top of my head can’t think of any other modality that has the tools and the information that we have to work with women. We have the tools to help them address every aspect of that [preconception health] really, and obviously we have those people to refer out to for the things that we don’t, but we can help a person to understand fundamentally what it means to be a healthy human person, which then puts that person in the optimal position to have a healthy reproductive system.” (Naturopath 7)*

The midwives in our study felt that they, too, have a valuable role but feel overlooked for their contribution to providing preconception care services, as expressed by this midwife:

> *“So, it is quite important, more important than anybody really realised, and I think as midwives, we are undervalued and not used as we could be for these sorts of things.” (Midwife 4)*

The health professionals in our study shared their views on other health professionals who could be referred to for preconception care. These included health professions from medical specialists (i.e., geneticists, obstetricians and gynaecologists, psychiatrists, neurologists, gastrologists, fertility specialists, endocrinologists) and allied health care professionals (i.e., dentists, acupuncturists, physiotherapists, social workers or counsellors, pharmacists, diabetes educators, exercise physiologists, pathologists, massage therapists, osteopaths, psychologists, and dieticians).

### Health professionals other than GPs want greater coordination of preconception care

Both midwives and naturopaths in our study describe wanting greater coordination of care with GPs to provide preconception care within the community. Both naturopaths and midwives shared their view that there is a need to educate GPs on the potential role they can provide in preconception care and where there are referral opportunities as described by this naturopath and midwife:

> *“I think we have to have a confidence in what we do well and work with as many GPs as we can to educate them on what we do and what we’re able to do, that they don’t have time, perhaps they’d appreciate more of what we can do. And some certainly do, and we do get referrals in from GPs.” (Naturopath 4)*

> *“Do we need to also help educate our GPs on our value as midwives and to improve our value within the primary health care setting?” (Midwife 3)*

Naturopath participants also shared some frustration regarding their experience of coordinating care with GPs for preconception care for their patients. As expressed by these two naturopaths the central issue raised was a perceived tension between the health professions when making requests of the GP concerning the care of their mutual patient:

> *“My relationship with some GPs is excellent, and with others you know that they’re a little bit more tricky to deal with. We do try and write to all of the GPs, and certainly, if we’re suggesting they go back to the GP to see the GP, we’ll extend the tests. It’s a very tricky position, as you know, with us not having any Medicare rebate for pathology testing, and how we straddle that. So, we do do a lot of referring for pathology direct, so that the patient doesn’t pay out-of-pocket, but in some instances I think it’s indicated for the doctor to consider other test, and then it’s writing I hope, a professional letter that doesn’t get their backup.” (Naturopath 4)*

> *“I would like to see the mainstream medical profession, naturopathy, all these natural therapies and mainstream medicine working hand in hand, and to stop this war that’s going on, we could do it better in terms of the medical profession because we all need to do it together. It’s not a competition.” (Naturopath 6)*

## Discussion

While the findings from this study suggest a range of limiting factors to the delivery of effective preconception care services within Australian primary health care settings, future preconception care opportunities were also identified.

The health professionals in our study identified the importance of making the public more aware of preconception health outside of clinical settings and would like to see an increase in public preconception health awareness and education initiatives across the life course. Other studies have reported the lack of patient awareness of preconception care as a potential barrier to preconception care [28-30]. Aligning clinical care and public health initiatives is important. Proposed strategic models for addressing reproductive health needs across the life course [31] include integrating clinical-based care, including preconception care with reproductive planning, preconception health school-based education, social media, and national campaigns [31]. Preconception health promotion messages tailored to people’s motivation and readiness for information across the reproductive life course are proposed as effective strategies [32]. To support clinicians in providing preconception care, more focus and clearer public health policy for the implementation and evaluation of proposed strategic models of healthcare that address preconception health across the reproductive life course for both women and men is needed.

Furthermore, as highlighted by the findings in our study, gender bias towards women needs to be addressed in preconception health education initiatives so as not to burden women with sole responsibility or to place a barrier to men receiving preconception care [33]. A lack of awareness regarding the role of men in preconception care is an issue described in other studies [19, 33, 34]. Research is limited that describes the male perspective regarding informed decision-making concerning fertility and reproduction [35-37]. Some of the underlying reasons for men’s lack of involvement are the perception by health professionals, women and men that when it comes to fertility and reproductive health, this is a women’s domain [33]. Men generally demonstrate poor knowledge of fertility factors yet report wanting to be involved in childbearing discussions and to improve their knowledge of fertility [38]. A recent systematic review of the literature (n=11 studies) on men’s knowledge of preconception health suggests that men’s preconception health knowledge is low [39]. Health professionals, researchers, and policymakers need to proactively encourage male involvement in reproductive decision-making [33]. To achieve this, male-inclusive healthcare services need to be co-designed with males and made widely available, while education for preconception health should also target adolescent and young adult males [33]. There are gender differences in health literacy that should also be considered by health professionals when providing preconception health information to men [40, 41]. In other studies, health professionals relate their confidence in providing preconception care to their level of preconception health knowledge, including men’s preconception health [19, 20, 42, 43], emphasising the importance of education for health professionals. Health professionals should think more carefully about engaging with men in preconception care. More training and patient information resources are needed to support primary care clinicians providing preconception care to men.

Our findings highlight barriers to formal coordination of preconception care between health professionals in Australian primary care health settings. Other studies also report a lack of referral and coordination between health professionals as a barrier to preconception care [28, 30, 43, 44]. In this study, the call for greater coordination of care comes from naturopaths and midwives, not GPs. While GPs are positioned in the health system as responsible for the coordination of care[45], there are potential contributions to preconception care that midwives and naturopaths believe they can make due to their position in the health system, their skills and the populations serviced. Midwives believe they have a responsibility to provide preconception care [30] in the postpartum period, and evidence suggests they are uniquely positioned to enquire about a woman’s desire for future pregnancy with opportunities to offer interconception care [46]. Midwives are providers of health counselling and education that includes a broad range from antenatal, preparation for parenthood, women’s health and sexual and reproductive health care [47]. In the current Australian healthcare model, midwives report limitations in being able to practice to their full scope of practice, including constraints on public healthcare funding to support contemporary midwifery practice [47]. Naturopaths are also accessed by women wanting to conceive. They also provide preconception care [14, 15] and other healthcare services aligned with preconception care, including addressing health risks and behaviours such as dietary practices, physical activity, alcohol consumption and illicit drug use [22]. Naturopaths report wanting greater collaboration and integration with other health professionals [22]; however, in Australia, naturopaths report facing barriers to collaborating with other health professions due, in part, to their exclusion from the National Registration and Accreditation Scheme [48], and other health professionals’ low confidence in or understanding of the efficacy and safety of naturopathic practices in Australia [49]. Future preconception healthcare policy and workforce planning need to consider the potential contribution of these and other health professions to meet the preconception care needs of the community rather than relying solely on the overburdened GP services. An integrated and collaborative whole health system approach to preconception care is needed and may provide more opportunities to deliver preconception care to improve reproductive health outcomes.

## Limitations and Future Directions

This study presents original findings from three primary care professions involved in preconception care in Australia. Although, there are some limitations to his study. The study findings may reflect characteristics specific to the Australian health system and context as only Australian health professionals were sampled. The focus groups were at the minimum sizes ideal for focus group dynamics [50, 51]. Notwithstanding these limitations, the study still identifies novel insights that warrant further exploration. Due to the nature of qualitative research and the self-selection of patients, the data should not be seen as generalisable. Further investigation should be conducted through quantitative research (e.g., survey) to identify self-reported practice behaviours in the broader community of these primary care professions. Third-party observational studies can be employed to triangulate health professionals’ self-reported practice behaviours concerning preconception care.

## Conclusions

This study provides key insights into health professionals’ limitations and scope of preconception care within Australian primary care settings as viewed by primary care practitioners. Health professionals have identified that making the public more aware of preconception health and aligning clinical care and public health initiatives are important. Health professionals should think more carefully about engaging with men in preconception care. Encouraging men to participate in preconception care requires different information and approaches. While GPs are positioned in the healthcare system as responsible for the coordination of care, there are potential contributions to preconception care that other health professionals can make due to their position in the healthcare system, their skills and the populations they service. A complementary and collaborative whole healthcare system approach to preconception care is needed and may provide more opportunities to deliver preconception care to improve reproductive outcomes.

## Supporting information

Additional file1. Focus group and interview guide

## Data Availability

The datasets used and/or analysed during the current study are available from the corresponding author upon reasonable request.

## List of abbreviations

ACM: Australian College of Midwives
CMA: Complementary Medicines Association
GP: General practitioner
NHAA: Naturopaths and Herbalists Association of Australia
RACGP: Royal Australian College of General Practitioners

## Declarations

### Ethical approval and consent to participate

The research was approved by the University of Technology Sydney Human Research Ethics Committee (ETH20-5547).

### Consent for publication

Not applicable

### Author contributions

The authors confirm their contribution to the paper: study conception and design: CC, AS; data collection: CC, AS; analysis and interpretation of results: CC, AS; draft manuscript preparation: CC, AS, DS, EM. All authors reviewed the results and approved the final version of the manuscript.

## Acknowledgements

The authors thank member associations RACGP, RACGP Specific Interest group Antenatal and Postnatal Care, ACM, NHAA and CMA representing GPs, midwives, and naturopaths for supporting this research by promoting the study to their members for participation. The authors also thank each health professional who gave their time to participate in the study. The authors thank Endeavour College of Natural Health for supporting the research study by awarding a project grant.

## Additional files

Additional file 1. Focus group and interview guide

